# Assessing the Impact of Mental Health Difficulties on Young People’s Daily Lives: Protocol for a Scoping Umbrella Review of Measurement Instruments

**DOI:** 10.1101/2021.06.18.21259112

**Authors:** Karolin Rose Krause, Sophie Chung, Terri Rodak, Kristin Cleverley, Nancy J Butcher, Peter Szatmari

**Affiliations:** Cundill Centre for Child and Youth Depression, Centre for Addiction and Mental Health, Toronto, ON Canada; Clinical, Educational and Health Psychology, University College London, London, UK; Independent Health Researcher, London, United Kingdom; Centre for Addiction and Mental Health, Toronto, ON Canada; Lawrence Bloomberg Faculty of Nursing and Department of Psychiatry, University of Toronto, ON Canada; Department of Psychiatry, University of Toronto, ON Canada; Child Health Evaluative Sciences, Hospital for Sick Children Research Institute, Toronto, ON Canada; The Hospital for Sick Children, Toronto, ON, Canada

**Keywords:** Review, outcome measure, functioning, health-related quality of life, children, adolescents, mental health

## Abstract

**Introduction:** An important consideration for determining the severity of mental health symptoms is their impact on youth’s daily lives. Those wishing to assess life impact face several challenges: First, various measurement instruments are available, including of global functioning, health-related quality of life (HRQoL), and well-being. Existing reviews have tended to focus on one of these domains; consequently, a comprehensive overview is lacking. Second, the extent to which such instruments truly capture distinct concepts is unclear. Third, many available scales conflate symptoms and their impact, thus undermining much needed analyses of associations between the two.

**Methods and analysis:** A scoping umbrella review will examine existing reviews of life impact measures for use with 6-24-year-olds in the context of mental health and well-being research. We will systematically search five bibliographic databases (MEDLINE, Embase, APA PsycINFO, CINAHL, Web of Science), and conduct systematic record screening, data extraction and charting based on methodological guidance by the Joanna Briggs Institute (JBI). Data synthesis will involve the tabulation of scale characteristics, feasibility, and measurement properties, and the use of summary statistics to synthesize how these instruments operationalize life impact.

**Ethics and dissemination:** This study will provide a comprehensive road map for researchers and clinicians seeking to assess life impact in youth mental health, providing guidance in navigating available measurement options. We will seek to publish the findings in a leading peer-reviewed journal in the field. Formal research ethics approval will not be required.

**Registration details:** This protocol was registered prospectively with the Open Science Framework (osf.io/jfqdv).

**Strengths and Limitations of this Study:**

- Umbrella review methodology will enable a higher-level synthesis of existing review efforts, thus generating a comprehensive map of available measurement instruments and research findings about their properties.
- Our methodological approach is based on the JBI guidelines for scoping reviews, umbrella reviews, and psychometric reviews.
- This review is based on a rigorous systematic search developed and executed by a health science librarian.
- We will only include studies published in the English language since 1990.
- As this is a scoping umbrella review designed to map available measurement instruments, the quality and risk of bias of included review articles will not be systematically assessed.

## Introduction

A key consideration for determining the severity of mental health difficulties is the extent to which these difficulties impact on a young person’s daily life. The Diagnostic and Statistical Manual of Mental Disorders (DSM-5) [1] determines “clinical significance” in relation to two criteria: individuals must display specific symptoms, and those symptoms must cause considerable distress or impairment in daily life [2]. Impaired daily functioning has been shown to influence help-seeking and health providers’ decisions about the type of care an individual should receive [3,4]. Assessing life impact can also help contextualize changes in symptom severity scores when assessing treatment efficacy and effectiveness [5–10]. From a public health perspective, consideration of life impact has moved common mental health conditions like depression to the fore of public health agendas, by showcasing that the associated burden of disease is comparable to that of cardiovascular or respiratory diseases [2].

In child and youth mental health^1^ (hereafter we will use the terms “youth mental health” and “young people”), life impact has typically been approached through the lens of functional impairment [11–13]. Functioning describes a young person’s ability “to adapt to varying demands of home, school, peer group, or neighborhood” in line with age-specific expectations and cultural norms (p. 1060) [11]. On a continuum of functioning, impairment marks one end of the spectrum, while high levels of adaptation and competency (e.g., thriving, flourishing) mark the other end.

The Children’s Global Assessment Scale (CGAS) [14] is a commonly used single-item measure that provides an overall rating of a young person’s functioning, based on clinician report. Other instruments take a more fine-grained approach by assessing functioning in specific areas of life. For example, the Social Adjustment Scale [15] generates separate subscale scores for social functioning with friends, family, at school, and in dating contexts. In addition, measures of symptom- or condition-specific impairment, focus on the extent to which psychopathological “symptoms interfere with and reduce adequate performance of important and desired aspects of a child’s life” (p. 455) [16]. For example, the Strengths and Difficulties Questionnaire (Impact Supplement) [17,18], and diagnostic interviews like the Kiddie Schedule for Affective Disorders and Schizophrenia (K-SADS) [19] enquire about functional impairment caused by psychopathology symptoms indicated during earlier parts of the respective assessment.

In physical health contexts and some population-based research, the impact of a particular health condition or of a person’s overall health status on their daily life is often conceptualized as Health-Related Quality of Life (HRQoL). Quality of life, has been described as “the overall positivity with which individuals view their state and circumstances” (p. 455) [16], and is thought to span physical, mental and social well-being [16,20]. HRQoL refers more specifically to quality of life in a health or medical context [21]. Relevant instruments include, for example, the brief EuroQol 5D-youth that is commonly used in economic evaluations [22]; the 52-item KIDSCREEN that was developed for the measurement of HRQoL in the general paediatric population [23], or the PROMIS item bank for paediatric global health, designed to assess overall perceptions of health in youth with chronic health conditions [24].

Wellbeing is another domain that researchers may consider when assessing the life impact of mental health conditions. While a consensus definition is lacking, this domain has been described as “a combination of positive emotions, engagement, meaningful relationships, and a sense of accomplishment, or as flourishing in aspects of feeling and functioning, thus reflecting the positive aspects of mental health” (p. 771) [25]. For example, the Warwick-Edinburgh Mental Well-being Scale (WEMWBS) [26] is a self-report instrument validated in adolescents that exclusively assesses positive aspects of mental health.

The conceptual domains of functioning, HRQoL and well-being have different theoretical roots, yet it has been suggested these terms are often used interchangeably [16]. All three might be considered as avenues for assessing the life impact of mental health difficulties in children and youth. For example, a recently developed core outcome set for child and youth anxiety and depression recommends assessing functioning via three measures: the CGAS as a measure of clinician-rated global functioning; a self-report scale of condition-specific impairment; and the KIDSCREEN as an HRQoL measure [9]. More generally, it is not clear whether scales purported to assess life impact via these domains are truly conceptually distinct, or whether they merely focus on different ends of the functioning continuum [e.g., 27]. A systematic review of measurement instruments that examines degrees of overlap and complementarity is lacking.

Researchers wishing to assess the life impact of mental health difficulties in young people further face the challenge of selecting the most appropriate instrument. A recent scoping review identified 14 different measures of global functioning, three measures of condition-specific impairment, and 14 measures of health-related quality of life (HRQoL), across 257 treatment studies for child and youth anxiety, depression, obsessive-compulsive disorder (OCD) and post-traumatic stress disorder (PTSD) [9]. Several reviews provide overviews of available instruments [28–34], their measurement properties and feasibility characteristics, but these have tended to be domain-specific (e.g., focusing only on HRQoL); consequently, a comprehensive overview of life impact measures is lacking. On the other hand, broader reviews of mental health assessment tools [e.g., 35–37] have not typically been exhaustive in their coverage of life impact measures, and have not tended to examine methodological questions specific to life impact assessment.

A third challenge to the measurement of life impact in mental health is that many available instruments conflate items that assess symptom severity with items assessing the life impact of such symptoms. For example, the CGAS’s description of “superior functioning” includes “no symptoms” as a criterion [14]. Similarly, the Health of the Nation Outcome Rating Scale (HoNOSCA) [38] is a 13-item measure that includes seven symptom-focused items alongside five functional items (covering school functioning, self-care, and relationships with peers and at home). The conflation of symptom severity and life impact items in a single scale hinders analyses of cross-sectional and longitudinal associations between the two domains [16].

Finally, an important fourth challenge is that many available instruments have been developed in Western high-income countries, and may not have cross-cultural validity or measurement invariance in lower- or middle-income contexts or in specific cultural communities [e.g., 39]. As functioning is defined in relation to age-and culture-specific expectations and norms [11], life impact measures that are not culturally sensitive and appropriate may yield misleading data. Even in the contexts where measurement instruments were originally developed, youth may not always have been involved in their creation, which may weaken their content validity [40].

### Objectives and Research Questions

This scoping umbrella review will examine how functioning, impairment, HRQoL, and well-being have been conceptualized and measured in the child and youth mental health literature. It will seek to provide an overview of the design characteristics, feasibility aspects, and measurement properties of available instruments, by considering existing individual reviews as primary studies. We seek to answer the following research questions:

RQ 1. What child-, parent-, and clinician-reported measurement scales are available for assessing life impact in children and youth aged 6 to 24 years in the context of mental health and well-being research?

RQ 2. What information is available from existing reviews about the design characteristics (e.g., target construct, target age range and use context, intended informant), feasibility (i.e., length, cost and accessibility, language version availability), and measurement properties (i.e., validity, reliability, responsiveness) of these instruments?

RQ 3. What populations and use contexts were these instruments originally designed for, according to their initial development study? Which cultural contexts were the instruments validated in?

RQ 4. According to an instrument’s original development study, were young people consulted as part of the measure development process?

RQ 5. Do measures of functioning, HRQoL and well-being appear to capture distinct conceptual domains, as opposed to assessing the same domain at different ends of the functioning continuum, based on subscale and item content?

RQ 6. To what extent do available measures of life impact conflate the measurement of psychopathology symptoms with the measurement of life impact?

## Methods and Analysis

### Study design

The proposed study is a scoping umbrella review. An umbrella review considers existing review articles as its principal source of evidence and aims to compare, contrast, or synthesize their findings [41]. While systematic reviews (including umbrella reviews) typically seek to answer clearly defined questions (e.g., “Which measure of global functioning provides the highest degree of validity and reliability”), scoping reviews typically seek to answer broader questions about the state of the evidence or about predominant methodological approaches in a given area [42]. A scoping umbrella review is an appropriate approach for this study because several existing reviews can be synthesized to provide a comprehensive mapping of available instruments and their properties. We will follow the methodological guidance provided by the Joanna Briggs Institute (JBI) for the conduct of umbrella reviews, scoping reviews, and reviews of measurement properties [43–45].

### Protocol

This review protocol complies with the Preferred Reporting Items for Systematic Reviews and Meta-Analysis for Protocols (PRISMA-P) reporting guidelines (Appendix 1) [46]. The final review will follow the Preferred Reporting Items for Systematic Reviews and Meta-Analysis extension for Scoping Reviews (PRISMA-ScR) [42]. The protocol preprint was registered prospectively with the Open Science Framework (OSF) on 26 May 2021 [47]. Upon registration and submission of the protocol, title and abstract screening was complete, but full text screening had not begun. Any important amendments to this protocol will be documented on the OSF registration page.

### Inclusion Criteria

This scoping umbrella review will consider systematic reviews, scoping reviews, rapid reviews, and narrative reviews that seek to provide an overview of available measurement scales to assess functioning, impairment, HRQoL or well-being. These may be reviews that systematically identify a range of measurement instruments, or reviews that synthesize the available literature for a single instrument. Inclusion criteria for reviews are defined to match the PICO components for systematic reviews of measurement properties (i.e., Population, Instruments, Construct, Outcome) in line with JBI guidelines [45]. The PICO criteria are summarized in Table 1.

**Table 1.** PICO Statement for Scoping Umbrella Review.

|  | **P (Population)** | **I (Instruments)** | **C (Constructs)** | **O (Outcomes)** |
| --- | --- | --- | --- | --- |
| Included | Children and adolescents aged 6–24 years, with a primary mental health condition/concern, subject to mental health assessment in general population, or in the context of assessing life impact in health contexts broadly speaking. | Youth—, parent, clinician, or external rater report<br>Initial assessment or outcome measure | Global functioning<br>Social functioning<br>Functional impairment<br>HRQoL<br>Well-being<br>Flourishing | Domain<br>Target age group<br>Reporter<br>Target use context<br>Length<br>Accessibility & cost<br>Measurement properties |
| Excluded | Ages 0–5 or 24+<br>Children and youth with intellectual disabilities or where mental health is a secondary concern to a primary physical condition; pure physical health context | Performance test;<br>biometric assessment | Language ability<br>Cognitive ability<br>Executive functioning<br>Symptom severity |  |

#### Population (P)

Review articles must have an explicit focus on measurement in middle childhood (defined here as starting at age 6 in line with proposed age group standards [48]), adolescence, and/or young adulthood (defined here as ending at age 24, in line with the United Nations’ definition of “youth” [49]). Studies with a majority focus on adults will not be considered, unless they include a separate appraisal of tools for a relevant paediatric age group. We will exclude reviews focused on early childhood (i.e., ages 0–5 years), where tailored assessment approaches are likely needed.

We will include reviews that examine the measurement of life impact in populations with a primary mental health or substance use concern, or in the context of assessing mental health and well-being in the general population or in non-specific health contexts. We will exclude reviews that focus on youth with physical health conditions. Instruments identified in such reviews may place a considerable focus on physical body functions that may be less relevant in a mental health context. We will also exclude reviews focused on youth with intellectual disabilities, neurological conditions (e.g., epilepsy, cerebral palsy), or Autism Spectrum Disorder (ASD). These profiles may require specialized assessments of life impact, and the conceptual separation of symptoms from functioning may be particularly complicated (e.g., with social functioning constituting a symptom of ASD). A separate review may be warranted to cover life impact measures for children and youth with these conditions.

#### Instruments (I)

We will consider scales deigned for completion by clinicians, external raters, parents or carers, and young people. These may be assessment or outcome measures but must focus on an eligible domain of life impact (i.e., see below) rather than symptom severity or psychopathology. We will exclude reviews focused on diagnostic tools or on the assessment of specific mental health conditions, unless the article’s abstract explicitly states that life impact measures were considered alongside symptom severity measures. We will further exclude performance tests, cognitive tests, language assessments, biometric tests, school-based functional behavioural assessments [50] and population-level composite indices of well-being or HRQoL.

#### Constructs (C)

We will consider instruments designed to assess life impact through the measurement of global functioning, social functioning/adaptation, general or condition-specific impairment, HRQoL, well-being (including flourishing), and life satisfaction. Reviews are eligible if they are focused on the measurement of life impact, or if life impact domains are covered alongside other outcome domains (e.g., symptom severity). Constructs that are not eligible include symptoms of psychopathology, language ability, cognitive ability, executive functioning, and motor functioning. Instruments that cover any of these constructs at an item level as part of measuring a broader eligible construct (e.g., HRQoL) may be included.

#### Outcomes (O)

We will include articles that state an intent to review, appraise, or map relevant measurement instruments and that provide a structured discussion or a tabulated overview of the instruments identified.

#### Publication type

Reviews must have been published in the English language from January 1990 onwards. We will include review articles published in peer-reviewed journals, assessment handbooks (if accessible online), and conference proceedings (including workshop summaries and conference papers, but not including conference abstracts). We will further include reviews that were published as grey literature (e.g., as reports on organizational websites) and otherwise meet the inclusion criteria.

### Search Strategy

The development of the search strategy was led by a health science librarian (TR) in collaboration with other members of the review team (KKR, PS). The search strategy combines search terms describing the population (e.g., “child*” OR “youth” AND “depress* OR “anxiety disorder*” OR “externalizing problem*”) and domains of interest (e.g., “function* OR “HRQOL”) with search terms limiting the results to reviews (e.g., “systematic review” OR “scoping review”) of measurement instruments or outcome measurement approaches (e.g., “psychometr*” OR “measurement instrument*”). Our tailored search syntax is informed by existing search filters for measurement instruments that were developed by the University of Oxford’s Patient-Reported Outcome Measurement Group [51] and by the COnsensus-based Standards for the selection of health Measurement INstruments (COSMIN) initiative [52]. Pilot searches informed the final search strategy (see Appendix 2).

The final search was performed by the review team’s health science librarian (TR) in Medline, Embase, APA PsycInfo, Cumulative Index to Nursing & Allied Health Literature (CINAHL), and Web of Science, and by a member of the review team (KRK) in the COSMIN database of systematic reviews of outcome measurement instruments. Retrieved records were deduplicated using Covidence systematic review software [53].

We will ask a group of subject matter experts to review the list of articles identified through the automatic search, and to suggest additional reviews that may have been missed. We will also conduct a targeted grey literature search via specific databases and websites identified as relevant by the team’s health science librarian (TR). In addition, we will hand-search the reference lists of included reviews to identify and retrieve the original development papers associated with eligible instruments, as well as copies of the instruments themselves, as available. We will consider supplemental searches if key information about a measure’s design characteristics is not available from the identified reviews or the instruments’ original development studies. Due to resource constraints, we will not conduct supplemental searches for a measure’s feasibility characteristics or measurement properties, and will base our reporting for these aspects on the information available from existing reviews.

We will review clearinghouses of measurement instruments [35] for any additional scales that were missed by the included reviews, and will also make a note of any additional instruments identified while screening for eligibility. These additional instruments will not be subject to a systematic appraisal, but will be listed in the final report.

### Study Selection

Eligibility will be assessed via a two-stage screening process. For the title and abstract screening, 20% of all identified records were screened independently and in duplicate by two reviewers (KRK and SC). The kappa coefficient indicated substantial inter-rater agreement (*kappa* = 0.77) [54]. Discrepancies were discussed and a final inclusion or exclusion rating agreed. A single reviewer (KRK) then screened the remaining titles and abstracts. All records retained for full text screening will be checked for eligibility independently and in duplicate by two raters. Disagreements will be discussed and decisions about inclusion will be made with the help of a third reviewer as needed. Articles that do not meet inclusion criteria will be coded for exclusion in the Covidence software environment with the first exclusion criterion that becomes apparent. Eligible review articles will progress to data charting.

### Data Extraction and Charting

Data will be extracted and charted using tailored adaptations of the JBI data extraction templates for systematic reviews of measurement properties [45] and for umbrella reviews [44]. The adapted matrices will be piloted to ensure an appropriate level of detail is charted. Data extraction will be conducted by one review author, and spot checks for comprehensiveness and accuracy will be conducted on at least 20% of the included reviews by a second reviewer. Any disagreements will be resolved through discussion. Based on the extent of disagreement identified, the two reviewers will consider extending the spot checks to a larger subset of studies.

The information to be charted is shown in Table 2, below. We will refer to the original development studies as needed, to extract whether or not youth or families were involved in measure development. We will also extract in which contexts an instrument has been validated. Where possible, we may review each measure’s item content to indicate whether items cover symptoms of psychopathology as well as life impact, and to examine the extent of overlap between measures purported to assess different life impact domains. Depending on the number and accessibility of the instruments identified, we may seek to undertake a systematic item-level mapping of content [e.g., 55,56].

**Table 2.**
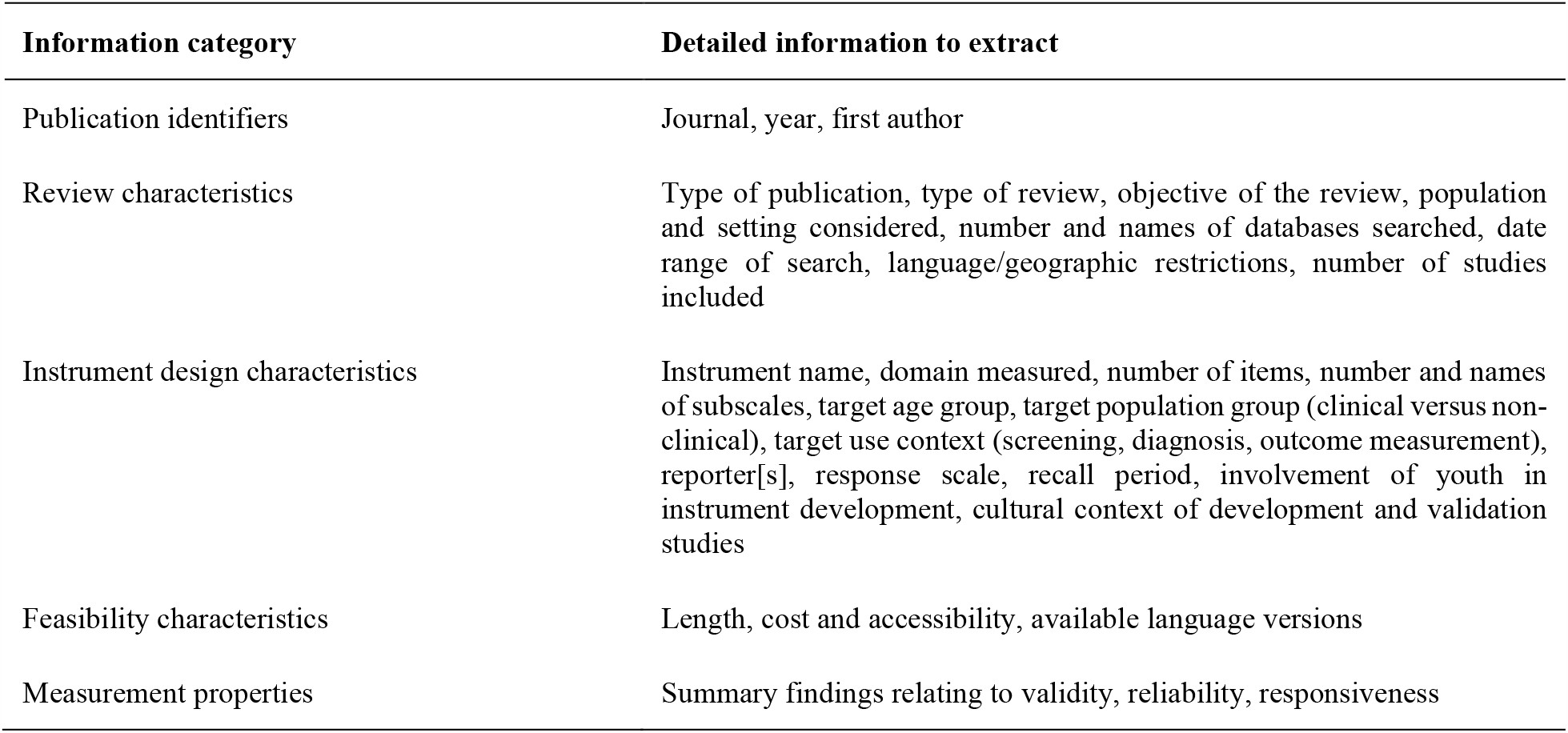
Overview of Date to Extract and Chart.

### Risk of Bias Assessment

Scoping-type reviews do not seek to generate critically appraised and summative responses to specific research questions, but instead aim to map the available evidence on a given topic. Therefore, risk of bias assessments are not typically conducted as part of scoping reviews [57], and are not planned for this scoping umbrella review.

### Strategy for Data Synthesis

We will synthesize the findings of existing reviews in relation to instrument design characteristics, feasibility characteristics, and measurement properties by applying the five-step data synthesis process recommended by Miles and Huberman [58] and Whittemore and Knafl [59]. This process consists of (1) data reduction; (2) data display; (3) data comparison; (4) conclusion drawing; and (5) verification. During verification, we will review the original development studies associated with each instrument to ensure that the information about key instrument characteristics compiled during the scoping umbrella review is accurate.

We will present the characteristics of the included review articles, as well as the characteristics of the identified measurement instruments in tabular format. We will report high-level quantitative summary statistics (i.e., counts or frequencies) to describe the reviews and instruments identified (e.g., number of instruments overall; number of instruments per life impact domain; number of instruments by type of reporter). We will further generate summary statistics and visualizations to report on the domains and subdomains of life impact covered by the identified instruments, and the extent to which these instruments appear to conflate items measuring symptoms of psychopathology with items measuring life impact, based on an examination of item or subscale content. We will also specifically indicate whether a measure was validated in a population with a mental health concern, or whether it was validated exclusively in community samples.

### Patient and Public Involvement

Draft review findings will be shared with a panel of youth advisors with lived experience of mental health challenges, and their feedback will be sought through a structured group discussion in order to help interpret and contextualize the review findings.

## Discussion

Historically, outcome measurement in youth mental health research has focused on symptom severity [60–62]. Yet, many common symptom scales are not immediately interpretable with regards to how a score change translates into real-world changes in a young person’s life. The assessment of life impact can provide important complementary information [5–7,63,64]. Two recent initiatives have highlighted functioning as a core outcome to track when evaluating clinical care for paediatric anxiety and depression [9], and when measuring youth mental health outcomes in population surveys [65]. Yet, difficulties have been reported with identifying a gold standard measure [9].

This scoping umbrella review does not aim to yield an authoritative summary of which instrument provides the best measurement properties. That would require an in-depth assessment of the methodological quality of the psychometric evidence underpinning each instrument in line with COSMIN guidelines [66], which in turn would constitute a study in its own right for each instrument identified [e.g., 67]. Instead, this review will examine a range of design, feasibility, and measurement properties to facilitate the pre-selection of candidates for in-depth psychometric appraisals. It further aims to identify gaps with regards to age groups or use contexts covered, and examine the degree of conceptual overlap between instruments designated to assess different outcome domains (e.g., functioning versus HRQoL). As such, it aims to take stock of current measurement practice, to inform discussions about suitable ways forward, and to provide a road map to researchers and clinicians seeking to appraise which tool or combination of tools may be appropriate for a given population and context.

### Dissemination

We will seek to publish the findings from this scoping umbrella review in a leading peer-reviewed journal in the field of child and adolescent mental health. We will also seek to disseminate findings at national and international conferences, and will consider submitting the final review to the COSMIN database of systematic reviews of measurement properties. We will also consider additional channels of dissemination, such as blog posts or podcasts.

## Supporting information

Appendix 1 PRISMA-P Checklist

Appendix 2 Medline Search Syntax

## Data Availability

This protocol includes no data.

## Footnotes

1 Defined here as spanning middle childhood (from 6 -11 years), early adolescence (12–18 years) and late adolescence (19–21 years), in line with the National Institute of Child Health and Human Development (NICHD Pediatric) Terminology [68]. We also include young adults up to the age of 24 years, in line with United Nations definitions [49].

