## Appendix 2 Medline Search Syntax for "Assessing the Impact of Mental Health Difficulties on Young People’s Daily Lives: Protocol for a Scoping Umbrella Review of Measurement Instruments"

**Search strategy for Ovid MEDLINE(R)**

1 mental health/

2 social adjustment/

3 mental disorders/ or exp anxiety disorders/ or exp "bipolar and related disorders"/ or exp "disruptive, impulse control, and conduct disorders"/ or exp dissociative disorders/ or exp "feeding and eating disorders"/ or exp mood disorders/ or exp "attention deficit and disruptive behavior disorders"/ or exp child behavior disorders/ or exp schizophrenia, childhood/ or exp neurotic disorders/ or exp personality disorders/ or exp "schizophrenia spectrum and other psychotic disorders"/ or exp somatoform disorders/ or exp substance-related disorders/ or exp "trauma and stressor related disorders"/

4 (mood disorder* or affective disorder* or personality disorder* or depressive disorder* or major depression or anxiety disorder* or obsessive compulsive or bipolar or schizo* or "substance use disorder*" or addiction* or conduct disorder* or behavio?r* disorder or stress disorder* or PTSD or CPTSD or complex trauma or developmental trauma).ti,ab,kf.

5 ((mental* or psychiatr* or internalizing or externalizing or psychosocial or psycho-social) adj3 (diagnos* or disorder* or ill or illness* or problem* or challenge* or issue* or difficult*)).ti,ab,kf.

6 (mental health or wellbeing or well-being).ti,kf.

7 or/1-6

8 (child* or pediatric* or paediatric* or youngster* or youth* or adolescen* or teen* or young adult* or young person* or young people* or juvenile* or student* or underage* or under-age* or emerging adult* or transition* age*).ti,kf,hw,jn.

9 Psychometrics/

10 "Outcome Assessment (Health Care)"/

11 Psychiatric Status Rating Scales/

12 "Surveys and Questionnaires"/

13 self report/

14 Mass Screening/

15 psychometr*.ti,ab,kf.

16 (clinimetr* or clinometr*).ti,ab,kf.

17 ((outcome* or rating or screening) adj2 (assessment* or scale*)).ti,kf.

18 measure*.ti,kf.

19 questionnaire*.ti,kf.

20 instrument*.ti,kf.

21 (("quality of life" or HRQL or HRQoL or QL or QoL or health profile* or health status* or global health or disabilit* or disabl* or function* or wellbeing or well being or flourish* or life satisfaction or impair* or interference or life impact* or adjust* or adapt* or standardi?ed or personali?ed or nomothetic or idiographic or self-report* or patient report* or child report* or youth report* or parent report* or clinician report* or mental health) adj3 (assess* or index or

indices or instrument or instruments or measur* or questionnaire* or profile* or scale* or score* or status* or survey* or apprais* or metric* or inventor* or tool* or indicator*)).ti,kf.

22 (health index* or health indices).ti,ab,kf,hw.

23 (PROM or PROMS).ti,ab,kf.

24 (HR-PRO or HRPRO).ti,ab,kf.

25 or/9-24

26 exp "Review Literature as Topic"/

27 "systematic review"/ (

28 meta-analysis/

29 (systematic review or scoping review or literature review or rapid review or narrative review or meta-analysis or metaanalysis).ti,ab,kf,hw,pt.

30 ("review of reviews" or "overview of reviews").ti,ab,kf.

31 ((review* or map*) adj5 (tool* or instrument* or scale* or measur*)).ti,ab,kf.

32 (literature adj3 review*).ti,ab,kf.

33 or/26-32

34 7 and 8 and 25 and 33

35 limit 34 to yr="1990 -Current"

***************************
